# Co-designing an online COmmunity suPporting familiEs after Sudden Cardiac Death (COPE-SCD) in the young

**DOI:** 10.1101/2021.05.20.21257559

**Authors:** Laura Yeates, Karen Gardner, Judy Do, Lieke van den Heuvel, Gabrielle Fleming, Christopher Semsarian, Alison McEwen, Leesa Adlard, Jodie Ingles

## Abstract

**Objective:** To co-design an online support intervention for families after sudden cardiac death (SCD) in the young (<35 years).

**Design:** Co-design of a SCD family intervention by stakeholder focus groups.

**Setting:** Families and healthcare professionals with experience in SCD in the young.

**Participants:** Semi-structured online focus groups were held with key stakeholders, i.e. family members who had experienced young SCD, healthcare professionals and researchers. Guided discussions were used to develop an online support intervention. Thematic analysis of discussions and iterative feedback on draft materials guided content development.

**Results:** Four focus groups were held (10-12 participants per group). Stakeholder involvement facilitated development of high-level ideas and priority issues. Creative content and materials were developed based on user preference for stories, narratives and information reflecting everyday experience of families navigating the legal and medical processes surrounding SCD, normalising and supporting grief responses in the context of family relationships, and fostering hope. Emphasis on accessibility led to the overarching need for digital information and online engagement. These insights allowed development of an online intervention - COPE-SCD: A COmmunity suPporting familiEs after Sudden Cardiac Death - which includes a website and online support program.

**Conclusion:** Using co-design with stakeholders we have developed a support intervention that directly addresses the needs of SCD families and fills a large gap in existing health care. We will evaluate COPE-SCD to determine whether this is an effective intervention for support of families following a young SCD.

**Strengths and limitations of this study:**

- Healthcare providers and consumer representatives participated as stakeholders in support intervention design.
- Co-design allowed development of a support intervention incorporating innovative ideas to meet user needs.
- Focus groups were limited in size and may not fully represent the needs of the wider community affected by sudden cardiac death in the young.

## INTRODUCTION

Sudden cardiac death (SCD) is a tragic complication of many genetic heart diseases, often occurring with no prior symptoms [1]. Causes of SCD can include inherited cardiomyopathies and inherited arrhythmia syndromes [2, 3]. These inherited conditions are mainly autosomal dominant, meaning first degree relatives are at a 1 in 2 (50%) risk of developing the same condition. They display clinical heterogeneity, with varying presentations from being asymptomatic through to heart failure and SCD.

Previous work has examined the psychological impact of SCD to the surviving family [4, 5]. Indeed, up to 1 in 2 family members report symptoms of posttraumatic stress and prolonged grief requiring referral to a clinical psychologist, on average six years after the death [5]. More recently McDonald *et al*. performed a needs analysis of parents who had lost a child to SCD, showing medical information and support as the most important needs, and psychological information and support (support from friends, family, community etc.) as the most unmet needs [6]. Further work by Steffen *et al*. identified four psychosocial needs to be addressed in future practice: a need for a safe environment, need to make sense of the death, need for affiliation and normalization, need to find new meaning and ongoing connection with the deceased [7]. Community or peer-based bereavement support groups can enhance social support, including an increased sense of wellbeing and personal growth among participants, and improved feelings of personal growth and positive meaning in life among peer providers [8].

Co-design is a collaborative change process that involves patients, family members, healthcare providers, researchers, and others in working together to design, develop and improve healthcare services. As demands on services continue to grow and become more complex, co-design is increasingly being recognised as an important method in health service improvement and as a key vehicle for delivering a more efficient and patient-focused system, [9] including for care of the elderly, the young, family services and chronic disease [11-14]. Involvement of end users in the deliberation and development of policy and programs will lead to more inventive ideas that better address user needs [10]. There is much to consider in improving support for families after SCD. Multidisciplinary care is needed, recognising the complexity of information to be shared and changing support needs that might be experienced over extended periods of time. We aimed to use co-design to develop an online support program for families affected by SCD in the young.

## METHODS

### Study design

Co-design was conducted over a series of steps (Figure 1). These involved (1) engaging stakeholders; exploring experiences, problems and current gaps in service delivery; (2) identifying new support options; and (3) refining and working to tailor these to the service context; following a similar process as previously described [15].

**Figure 1:**
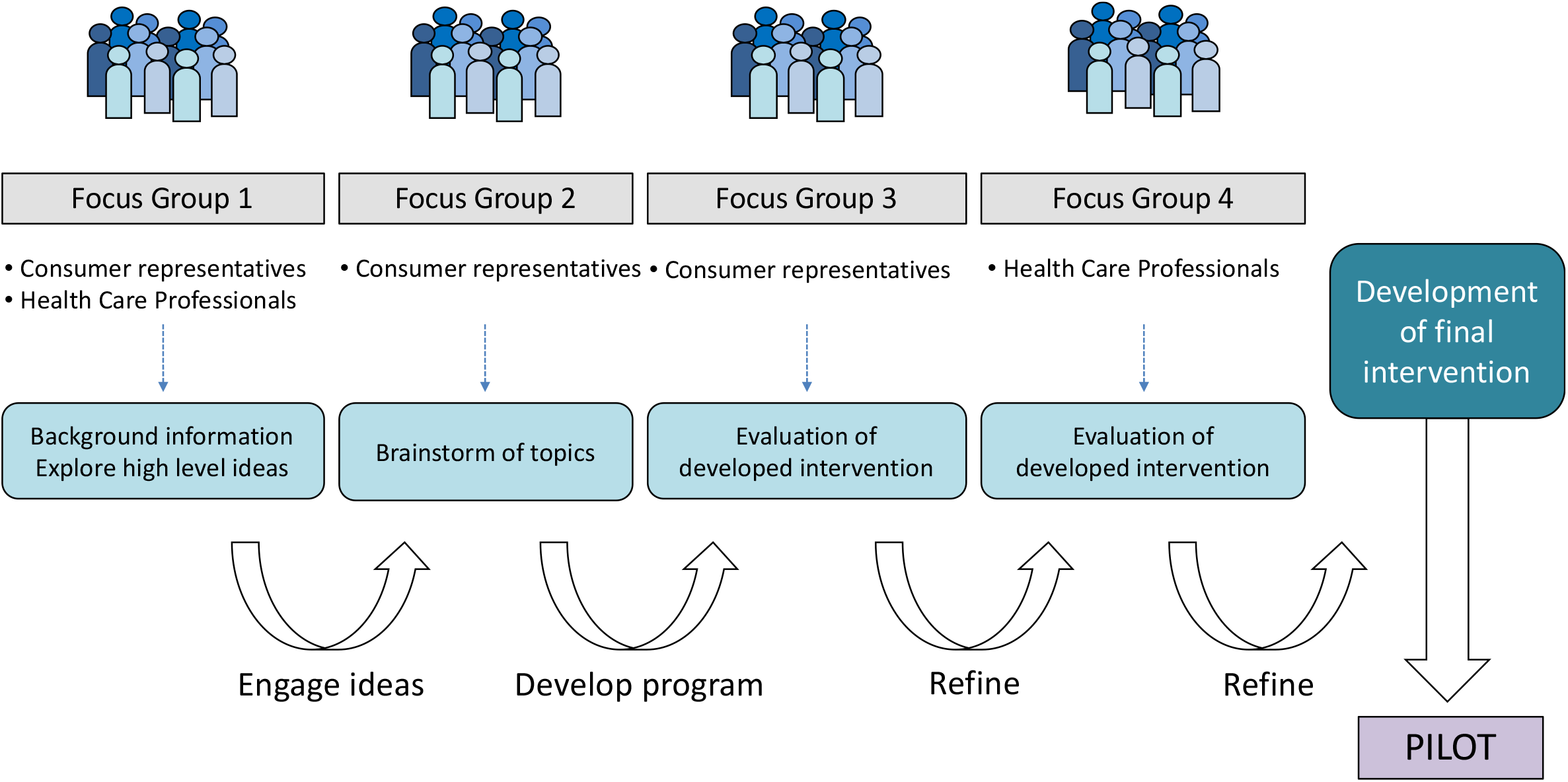
Evaluation of the intervention

### Consumer and stakeholder involvement

Purposive sampling was used to recruit key stakeholders including consumer representatives (i.e. family members who had experienced a young SCD), healthcare professionals with experience in caring for families after a SCD and researchers. Stakeholders were identified from a specialised genetic heart disease clinic (Sydney, Australia) and the service network in which it operates including forensic medicine departments, general clinical genetics departments, children’s hospitals and general cardiology.

### Patient and public involvement

Individuals with a family history of a SCD in a young first degree relative (consumer representatives) were invited to participate in stakeholder focus groups. Participants contributed ideas for a support intervention in focus groups 1 and 2 and provided feedback on the developed intervention in focus group 3 and 4.

### Focus groups

Guided discussion focus group sessions were held over video conference. These were recorded using Quicktime Player but not transcribed. The sessions were semi-structured and moderated by JI (genetic counsellor) with input from members of the research team, primarily LA (clinical psychologist), LY (genetic counsellor) as required. Discussion prompters e.g. Jamboard (jamboard.google.com) were used in some focus groups to organise material from discussions, enhance feedback on content of the intervention and guide content development. Each focus group had a different overall aim and specifically approached the stakeholder participants who could contribute to that aim (Figure 1).

### Data analysis and theme development

Field notes were taken (LY, JD) throughout each focus group session and when applicable, used in conjunction with discussion prompters to develop desired content themes for a support intervention.

### Prototype intervention development

Content themes (from focus group 2) were used as an outline in development of the proposed intervention which comprised two parts: a website, and an online support intervention. A draft intervention was then presented to focus group 3, comprising consumer representatives, where we invited discussion and feedback on the designed intervention. Between focus groups, the investigator team met periodically to discuss ideas generated during the co-design process. The intervention was further refined and presented to the final focus group 4 comprising healthcare professionals, for final feedback.

## RESULTS

Healthcare professionals were approached via email. Consumer representatives were approached by an initial phone call from LY, with follow-up information sent via email. Eighteen individuals (outside of the research team) were approached to participate in one or more focus groups. Of these, 11 participated in at least one focus group with 7 declining due to other commitments/ did not respond.

### Focus groups

Four focus groups were held (range: 10-12 participants per focus group) over a period of 4 months and comprised health care professionals, consumer representatives and members of the research team (Table 1).

**Table 1:** Characteristics of the focus group participants.

| **Participants n** | **Focus Group 1** | **Focus Group 2** | **Focus Group 3** | **Focus Group 4** |
| --- | --- | --- | --- | --- |
| **Consumer representatives** | **1** | **4** | **6** | **0** |
| **Healthcare professionals\*** | **8** | **4** | **3** | **9** |
| • Genetic Counsellor | 3 | 2 | 2 | 4 |
| • Cardiologist | 2 | 1 | 0 | 3 |
| • Clinical Psychologist | 2 | 1 | 1 | 1 |
| • Forensic Nurse | 1 | 0 | 0 | 1 |
| **Other participants** | **3** | **3** | **1** | **2** |
| • Peer researcher | 1 | 1 | 1 | 1 |
| • Research Assistant | 1 | 1 | 0 | 1 |
| • Student | 1 | 1 | 0 | 0 |
| **Total Participants** | **12** | **11** | **10** | **11** |

\*includes members of the research team

### Focus group 1 – Exploring high level ideas

The first focus group included both healthcare professionals and consumer representatives and introduced the rationale for the development of a support intervention. Participants then had the opportunity to engage in high-level discussion about their experiences and areas they felt needed to be addressed by a support intervention. Three key themes emerged from this session:

#### 1. The desire for a caseworker

Participants reported a “directionless” period following the sudden death of their family member and how a caseworker or a nominated contact would have provided them with general information as well as grief support. Healthcare professionals in this session, while supportive of this idea, acknowledged this would be an aspect of work needing significant commitment to funding.

#### 2. Gaps in the medical system

Participants recognised the limits of the current medical system and confusion about the referral pathways to access specialised multidisciplinary genetic heart disease clinics. Participants suggested a website could assist in providing accurate information for families and health care professionals after SCD, showing the pathways to finding appropriate specialists, and in addition may be a platform to allow families to connect.

#### 3. The need for peer support

Participants expressed a desire for peer support from families with shared and similar experiences. This was echoed by the healthcare professionals who recognised that appropriate support groups specific to SCD in the young could be a powerful mode of support. Participants expressed a preference for “something they can drop in and out of” acknowledging that their support and information needs varied at different times and easily accessible resources meant they could access as required.

### Focus Group 2 – Brainstorming content

The second focus group included consumer representatives and sought to brainstorm topics to be covered by a support intervention. Participants gave suggestions on content as well as different modes of delivery. Using discussion prompters and notes taken throughout the session, feedback was summarised into broad categories: Connecting, Information, Coping and Mode of delivery (Table 2). Participants gave suggestions for topics to be addressed, highlighting areas where information was difficult to access or where they experienced gaps in the care they received. This feedback was used to develop the intervention, ensuring topics raised were addressed where possible by multiple modes of the intervention recognising the need for easily accessed information in different formats.

**TABLE 2.** Summary of feedback from focus group 2, including examples and where these examples will be addressed in the intervention

| Content feedback | Overview | Examples | Addressed by COPE-SCD |
| --- | --- | --- | --- |
| **Connecting** | *Participants raised the desire to meet with other SCD families recognising the benefit of peer support* | Connecting with other families | Website & online session 1 |
|  |  | Tell your story | Website & online session 1 |
|  |  | Learning from the experience of others | Website & online sessions |
| **Information** | *Participants discussed the need for information, particularly around causes of death, coronial process & next steps for their family.* | Medical information: causes of sudden death | Website & online session 2 |
|  |  | Coronial process (expected timeline) | Website & online session 2 |
|  |  | Next steps/ plan for family | Website & online session 2 |
|  |  | Information for GP & health professionals | Website |
|  |  | Referral pathways | Website & online session 2 |
| **Coping** | *Participants sought help with coping after a sudden cardiac death* | Dealing with questions from family/ friends | Website & online session 3 |
|  |  | Returning to work/ school | Website & online session 3 |
|  |  | Coping with life | Website & online session 3 |
|  |  | How to talk about death | Website & online session 3 |
|  |  | Phases of grief | Website & online session 3 |
|  |  | Fear of repeat event | Website & online session 2 |
|  | *Participants sought help with coping in the context of the wider family* | Helping your family/ children grieve | Website & online session 4 |
|  |  | Different grief processes | Website & online session 3 & 4 |
|  |  | Addressing inaccurate assumptions of family/ friends/ community | Website & online session 4 |
|  |  | Reaching milestones: birthday, anniversary, other family events | Website & online session 4 |
|  |  | Coping as a family | Website & online session 4 |
|  |  | Family communication | Website & online session 4 |
| **Mode of delivery** | *Participants discussed the different ways they would like information presented* | Question & Answer sessions | All online sessions |
|  |  | Interactive | Online sessions |
|  |  | Personal Stories | Website & online session 1 |
|  |  | Written information | Website |
|  |  | Other information formats e.g. video | Website |
|  |  | Sessions for different family members (siblings, parents) | Online sessions |
|  |  | Practical help | Website & all online sessions |
|  |  | Support information for wider community | Website |

Further structuring of content topics were collated and grouped into three key themes (Figure 2):

1. **Uncertainty** – Including coping with uncertainty due to the cause of the SCD, uncertainty in the next steps for the participant and their family, the chance of reoccurrence of an SCD event in another family member.
2. **Individual coping –** including normalising different grief responses, coping with general life and returning to work/vocation.
3. **Family and systems coping –** understanding how people grieve differently within the family unit, change in family dynamics, practical support.

**Figure 2:**
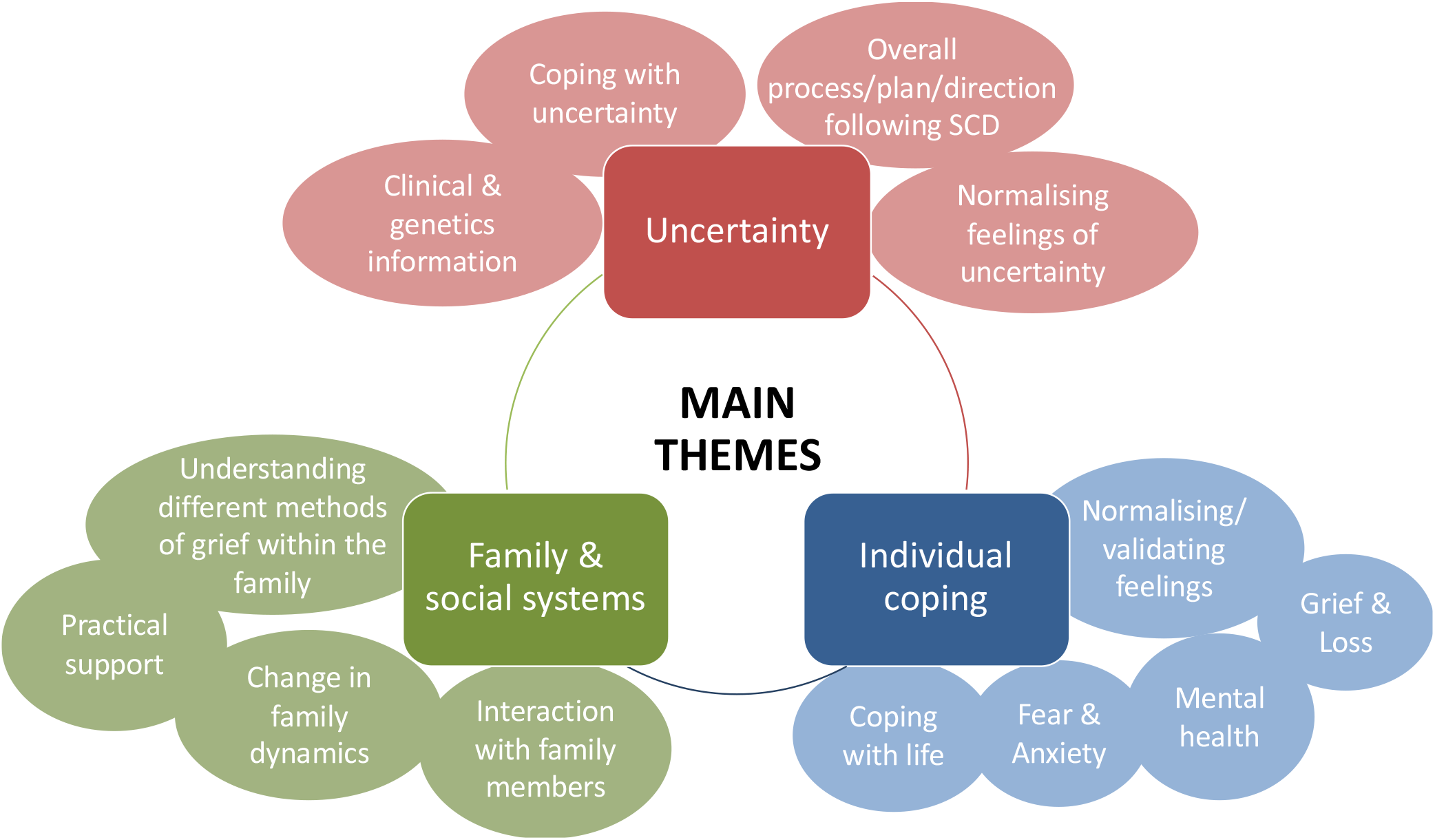
Key themes derived from the focus groups

### Focus Group 3 and 4 – Evaluation of proposed intervention

In the final two focus groups, the proposed intervention was presented to the group comprising consumer representatives (focus group 3) and health care professionals (focus group 4), inviting further feedback. Both groups gave constructive feedback in the overall content of the proposed intervention and the layout and structure, allowing further refinement of the interventions after each focus group.

### The support intervention: COPE-SCD - An online COmmunity suPporting familiEs after Sudden Cardiac Death in the young

The support intervention comprises two parts, a website and “online support sessions” (Figure 3). The website provides written and video content for families after a SCD. It contains both information on the causes of SCD, practical processes including the coronial process, as well as general information on grief and loss, individual and family coping and accessing further help. The website also has an area for family stories and information on the second part of the intervention, the online sessions.

**Figure 3:**
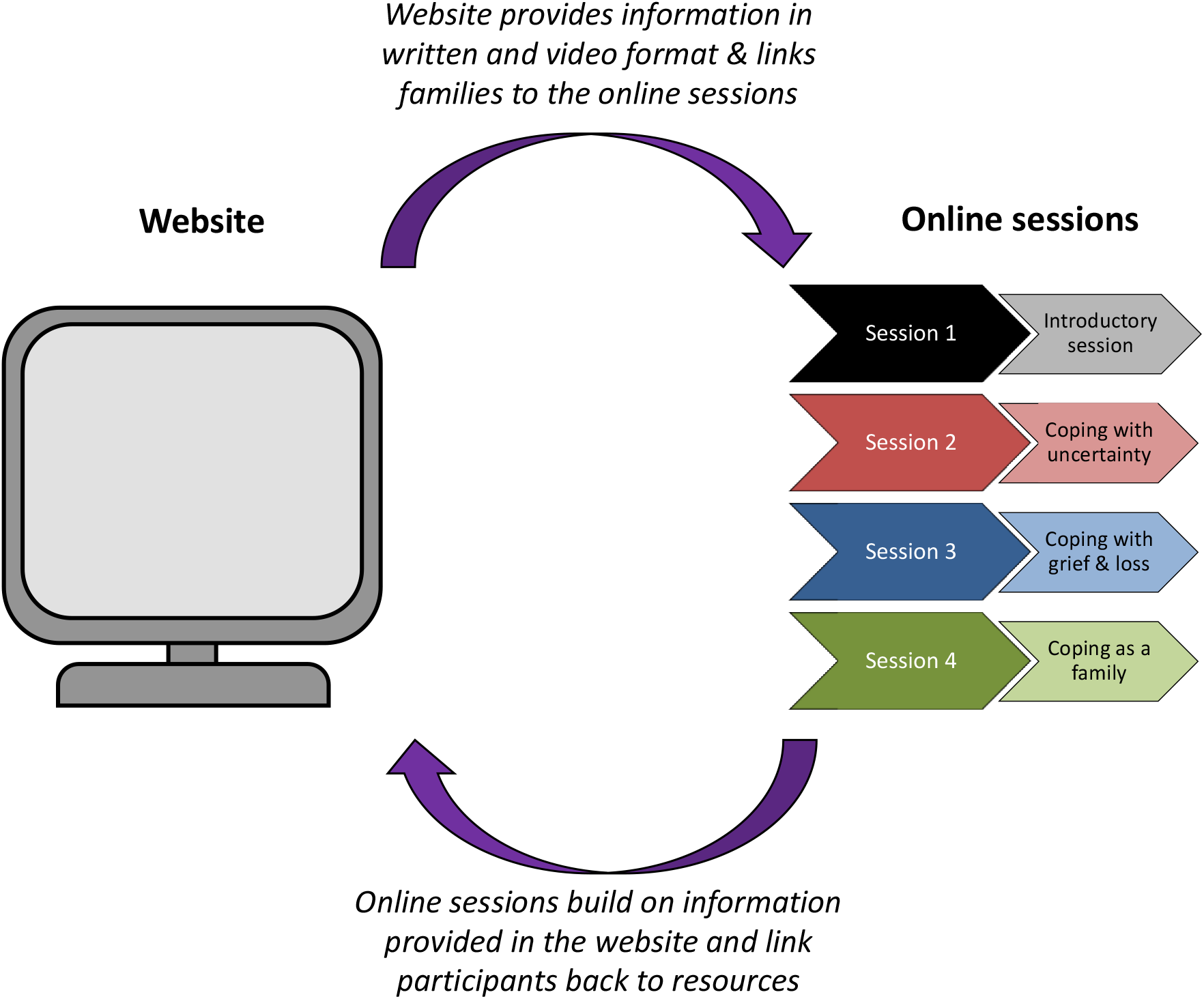
Overview of the COPE-SCD support intervention

The online sessions are a series of four live, virtual sessions held over video conference and facilitated by a clinical psychologist and genetic counsellor, with input from other health professionals. Each session has a specific focus (Figure 3) and provides a space for peer support and expert input from healthcare professionals. The sessions are interactive, encouraging participant input. Participants register for a group and the same group of individuals attend each of the 4 sessions through to completion.

The website and support sessions are complimentary aspects of the intervention, and allow for flexibility of access, acknowledging that the benefit of joining an online session group and the timing of such a group will differ from individual to individual. The combined intervention incorporates the consumer stakeholder priorities and their preference for stories, narratives and information reflecting everyday experience of families navigating the legal, medical, social and psychological processes surrounding SCD.

## DISCUSSION

Co-design is a method that promotes the preference of the end users by engaging them in the development process. We report our experience in using a co-design approach to develop a support intervention for families affected by SCD in the young. By engaging consumer representatives and health care professionals as stakeholders, we were able to shape and develop a support intervention, that aims to fill a gap in clinical care and directly address user needs.

Co-design facilitated the identification of key priority areas for a support intervention and highlighted areas of greatest need, allowing their prioritisation into the designed intervention. Involving both healthcare professionals and consumer representatives, enabled the process to identify gaps and problems in the current care system provided to SCD families, from the perspective of both the recipients and health care providers. This approach gave wider scope to design a program that both fulfills the needs of families affected by SCD, whilst ensuring a practical and complimentary addition to current services.

Peer support specific to SCD was identified by participants as a significant gap in current support services. Peer support is a recognised intervention across health care and has been shown to be of benefit to adults after the death of a family member [8]. Peer support is underpinned by the premise that supportive interactions with people who have experienced similar difficulties can give individuals a sense of empowerment, increase self-efficacy, and enhance coping skills [16]. Peer support programs come in many different forms and can be led by professionals or volunteer peers. Peer support volunteer led groups involve people with similar backgrounds providing emotional, social, or practical support to each other [17]. Peer supporters draw on their shared experiences to provide empathic understanding, information, and advice to those they are helping. A key aim is to promote hope, recovery from illness or trauma, improve life skills, psychological well-being, and social integration [18]. Professionally led groups are therapeutic in nature, and focus on developing treatment goals within a group setting [19]. They provide participants with both peer and professional support, encouraging sharing of experiences and feedback to facilitate greater insight and personal change [20]. Participants in this study raised the specific need of a shared experience of SCD, when they sought peer support. Recognising the impact of a previously well person dying suddenly with no apparent cause/ undiagnosed genetic heart disease, was very different to death by long-term illness or an external cause such as motor vehicle accident.

Including bereaved patient groups in research has been previously discussed, with many studies reporting bereaved research participants find therapeutic value in their research participation and are motivated by the desire to help others going through a similar situation [21]. In our study, the patient group had a long-standing relationship with the clinical-research team. The process of engaging end users was straightforward as long-term trusting relationships had already been established over many years. The desire to meet and help others in a similar situation appears to be strong motivator in this unique group and established trust relationships made it possible to provide a co-design space for open and honest discussion. While various levels of co-design have been described in the literature, from involving end users in the design phase to involving them in the delivery [22], this co-design process was focused on design. Some participants expressed a view that involving peers in delivering support programs would also be a welcome addition to care. Further discussion of such options may be addressed as the program develops, is tested and becomes established. A family affected by SCD will encounter a number of health care professionals as they seek to determine the cause of death of their family member. While referral to key services such as a specialised multidisciplinary genetic heart disease clinic, support services and connection with research centres seems a straight forward process, participants described this as confusing, slow and difficult. Navigating the legal and medical processes surrounding SCD, normalising and supporting grief responses in the context of family relationships, and fostering hope were key priorities for SCD families, yet they often describe the difficulty in accessing services. Variability in access to service was experienced across residential locations, where those out of major cities had greater difficulty accessing specialised clinics, information and support. Thus, the aim of COPE-SCD is to provide a centralised source of information that connects a family who has experienced SCD with other families, and with the appropriate services. Online delivery means there can be access to information that can be revisited as needed and supports equitable access for those in rural and remote areas. Coupled with this is the peer support component which comprises family stories housed on the website and access to the online sessions. COPE-SCD aims to provide families and communities with resources and support, and fills an existing gap in the care of families after SCD.

Co-design was incorporated into the development of this support intervention with relative ease. The motivation of the stakeholder groups, including both consumers and healthcare professionals, to engage and be involved in the research has produced, in a relatively short timeframe, a great depth of insight into the needs of families and their priorities for support. The result is an intervention that families believe will be of assistance in the most tragic of circumstances and one that will equip health care professionals with practical and helpful tools so they may provide better support.

## CONCLUSION

SCD in the young is a devasting event which has lifelong impact on the family and supporting community. Support interventions that better care for families following SCD are highly sought after and currently are an existing gap in clinical care. Using co-design with stakeholders we have developed a support intervention that directly addresses the needs of end users and fills a gap in existing health care. The service providers and end user group in this co-design process were highly motivated to participate and deep insights into the experiences of families following SCD have shaped the priorities and specific interventions developed. Next steps will include evaluation of the intervention.

## Data Availability

De-deidentified data are available at reasonable request.

## ACKNOWLEDGEMENTS

We would like to thank the focus group participants.

## CONTRIBUTORS

LY wrote the majority of the first version of this manuscript as a PhD student under the supervision of JI, CS and AM. KG wrote sections of the first version of the manuscript and assisted in supervision of the project. JI, LA, JD and LVH conceived the study design. LY collected field notes on all focus groups, JD collected field notes on focus group 1, 2 and 4. GF and AM were involved in thematic analysis of focus group 1. LY and JD conducted thematic analysis of focus group 2. LY, KG, LA, JD, LVH, CS and JI contributed to analysis of each focus group and development of the intervention between groups. JI was the senior author and oversaw the entire project. All authors reviewed and gave their approval for the final manuscript prior to publication.

## ETHICS APPROVAL

This study was approved by the Ethics Review Committee (RPAH Zone) of the Sydney Local Health District (protocol number X19-0405).

## DATA AVAILABILITY STATEMENT

De-deidentified data are available at reasonable request.

## REFERENCES

1. Stiles, M.K., et al., 2020 APHRS/HRS expert consensus statement on the investigation of decedents with sudden unexplained death and patients with sudden cardiac arrest, and of their families. Heart Rhythm, 2020.

2. Bagnall, R.D., et al., A Prospective Study of Sudden Cardiac Death among Children and Young Adults. N Engl J Med, 2016. 374(25): p. 2441–52.

3. Semsarian, C., J. Ingles, and A.A. Wilde, Sudden cardiac death in the young: the molecular autopsy and a practical approach to surviving relatives. Eur Heart J, 2015. 36(21): p. 1290–6.

4. Yeates, L., et al., Poor psychological wellbeing particularly in mothers following sudden cardiac death in the young. Eur J Cardiovasc Nurs, 2013. 12(5): p. 484–91.

5. Ingles, J., et al., Posttraumatic Stress and Prolonged Grief After the Sudden Cardiac Death of a Young Relative. JAMA Intern Med, 2016. 176(3): p. 402–5.

6. McDonald, K., et al., Needs analysis of parents following sudden cardiac death in the young. Open Heart, 2020. 7(2).

7. Steffen, E.M., L. Timotijevic, and A. Coyle, A qualitative analysis of psychosocial needs and support impacts in families affected by young sudden cardiac death: The role of community and peer support. Eur J Cardiovasc Nurs, 2020. 19(8): p. 681–690.

8. Bartone, P.T., et al., Peer Support Services for Bereaved Survivors: A Systematic Review. Omega (Westport), 2017. 80(1): p. 137–166.

9. Bate, P. and G. Robert, Experience-based design: from redesigning the system around the patient to co-designing services with the patient. Qual Saf Health Care, 2006. 15(5): p. 307–10.

10. Trischler, J., T. Dietrich, and S. Rundle-Thiele, Co-design: from expert-to user-driven ideas in public service design. Public Management Review, 2019. 21(11): p. 1595–1619.

11. Yadav, U.N., et al., Evaluating the feasibility and acceptability of a co-design approach to developing an integrated model of care for people with multi-morbid COPD in rural Nepal: a qualitative study. BMJ Open, 2021. 11(1): p. e045175.

12. Steen, M., M. Manschot, and N. De Koning, Benefits of Co-design in Service Design Projects. 2011.

13. Hjelmfors, L., et al., Using co-design to develop an intervention to improve communication about the heart failure trajectory and end-of-life care. BMC Palliat Care, 2018. 17(1): p. 85.

14. Banbury, A., et al., Adding value to remote monitoring: Co-design of a health literacy intervention for older people with chronic disease delivered by telehealth - The telehealth literacy project. Patient Educ Couns, 2020. 103(3): p. 597–606.

15. NSW Government and Agency of Clinical Innovation. A Guide to Build Co-design Capability. 2019; Available from: https://aci.health.nsw.gov.au/data/assets/pdf_file/0013/502240/Guide-Build-Codesign-Capability.pdf.

16. Pistrang, N., C. Barker, and K. Humphreys, Mutual help groups for mental health problems: a review of effectiveness studies. Am J Community Psychol, 2008. 42(1-2): p. 110–21.

17. Solomon, P., Peer support/peer provided services underlying processes, benefits, and critical ingredients. Psychiatr Rehabil J, 2004. 27(4): p. 392–401.

18. Landers, G.M. and M. Zhou, An analysis of relationships among peer support, psychiatric hospitalization, and crisis stabilization. Community Ment Health J, 2011. 47(1): p. 106–12.

19. Cohen, M.B. and A. Mullender, The Personal in the Political: Exploring the Group Work Continuum from Individual to Social Change Goals. Soical Work with Groups, 2005. 28(3-4): p. 187–204.

20. Mason, E.C. L; Pistrang, N, Processes and experiences of mutual support in professionally-led support groups for people with early-stage dementia. Dementia, 2005. 4(1): p. 87–112.

21. Beck, A.M. and C.A. Konnert, Ethical Issues in the Study of Bereavement: the Opinions of Bereaved Adults. Death Studies, 2007. 31(9): p. 783–799.

22. Co-production in social care: what it is and how to do it. 2015; Available from: https://www.scie.org.uk/publications/guides/guide51/.

